# Clinical testing panels for ALS: global distribution, consistency, and challenges

**DOI:** 10.1101/2022.09.27.22280431

**Authors:** Allison A. Dilliott, Ahmad Al Nasser, Marwa Elnageeb, Jennifer Fifita, Lyndal Henden, Ingrid M. Keseler, Steven Lenz, Heather Marriott, Emily McCann, Maysen Mesaros, Sarah Opie-Martin, Emma Owens, Brooke Palus, Justyne Ross, Zhanjun Wang, Hannah White, Ammar Al-Chalabi, Peter M. Andersen, Michael Benatar, Ian Blair, Johnathan Cooper-Knock, Luke Drury, Elizabeth Harrington, Jeannine Heckmann, John Landers, Cristiane Moreno, Melissa Nel, Evadnie Rampersaud, Jennifer Roggenbuck, Guy Rouleau, Bryan Traynor, Marka van Blitterswijk, Wouter van Rheenen, Jan Veldink, Jochen Weishaupt, Matthew B. Harms, Sali M.K. Farhan, the Amyotrophic lateral sclerosis spectrum disorders Gene Curation Expert Panel

## Abstract

**Objective:** In 2021, the Clinical Genome Resource (ClinGen) amyotrophic lateral sclerosis (ALS) spectrum disorders Gene Curation Expert Panel (GCEP) was established to evaluate the strength of evidence for genes previously reported to be associated with ALS. Through this endeavor, we will provide standardized guidance to laboratories on which genes should be included in clinical genetic testing panels for ALS. In this manuscript, we aimed to assess the heterogeneity in the current global landscape of clinical genetic testing for ALS.

**Methods:** We reviewed the National Institutes of Health (NIH) Genetic Testing Registry (GTR) and members of the ALS GCEP to source frequently used testing panels and compare the genes included on the tests.

**Results:** 14 clinical panels specific to ALS from 14 laboratories covered 4 to 54 genes. All panels report on *ANG, SOD1, TARDBP*, and *VAPB*; 50% included or offered the option of including *C9orf72* hexanucleotide repeat expansion (HRE) analysis. Of the 91 genes included in at least one of the panels, 40 (44.0%) were included on only a single panel. We could not find a direct link to ALS in the literature for 14 (15.4%) included genes.

**Conclusions:** The variability across the surveyed clinical genetic panels is concerning due to the possibility of reduced diagnostic yields in clinical practice and risk of a missed diagnoses for patients. Our results highlight the necessity for consensus regarding the appropriateness of gene inclusions in clinical genetic ALS tests to improve its application for patients living with ALS and their families.

## Introduction

Historically, for patient stratification and counseling purposes, ALS cases have been largely, and perhaps misleadingly, classified as familial (fALS) or sporadic (sALS), referring to the ∼10% of cases with a family history and ∼90% of cases without a family history, respectively. While family history may be a reasonable proxy for the expectation that a genetic cause of disease might be identified, it is increasingly clear that monogenic causes may also account for cases of sALS. Indeed, a Mendelian-inherited, monogenic cause for disease may be found in 40–55% of fALS cases, with variants in the genes *SOD1, FUS, TARDBP*, and *C9orf72* — specifically, the GGGGCC hexanucleotide repeat expansion (HRE) — being most common (1), but 5-10% of sALS cases have also been found to carry the *C9orf72* HRE (2-4). In fact, most, if not all, genetic causes of fALS have also been described in patients with apparently sporadic ALS, underscoring the importance of not conflating fALS with monogenic ALS.

Clinically, genetic testing offers many advantages, the most critical being early and accurate diagnosis. Due to the large amount of heterogeneity in clinical presentation, disease progression and prognosis, and the frequent presence of additional clinical features not related to the motor system, such as frontal temporal dementia (FTD) among ALS patients, misdiagnosis in early ALS onset is common (5, 6). However, genetic testing can end the diagnostic odyssey many patients experience, and there is particular interest in genetic testing due to the increasing number of clinical trials enrolling specific genetic populations (7). Ongoing trials include those for symptomatic ALS patients carrying pathogenic variants in *ATXN2, C9orf72, FUS*, and *SOD1* (NCT04494256, NCT03626012, NCT04931862, NCT04768972, NCT02623699, NCT05039099) (8). A recent prospective assessment of genetic testing in an ALS referral clinic suggested that routine screening for pathogenic variants in ALS patients may impact clinical care in at least 21% of cases (9). Moreover, ALS patients and their families are increasingly aware of the relevance of genetic testing for at-risk family members (4, 10, 11), and clinical trials for unaffected pre-symptomatic gene mutation-carrying family members have begun to emerge (NCT04856982) (12). Patient interest and the potential for impact on clinical care has resulted in an increase in the number of clinicians utilizing genetic testing for their ALS patients. A 2017 survey of 167 clinicians from 21 countries found that over 90% of respondents would offer genetic testing to their fALS patients, and 49% of respondents would offer genetic testing to their sALS patients (13), with similar results being obtained from a 2021 Canadian study (8). However, both studies also found a lack of consensus regarding which ALS patients should be eligible for genetic testing and the testing practices that should be used (8, 13).

A critical aspect of clinical genetic testing for ALS is the determination of which genes are assessed for pathogenic mutations driving disease. Typically, multi-gene clinical genetic panels are employed because of their efficiency and cost-effectiveness (11). A vast number of genes have now been implicated in ALS (14), with widely varying levels of evidence supporting the strength of the causal relationship. Yet, clinical testing laboratories have no standardized way of assessing the validity of gene-disease relationships and have made idiosyncratic decisions regarding which genes to include on testing panels. Concerningly, the variability of genes included could result in disparate diagnostic yields in clinical practice and risk of a missed diagnosis for individual patients; in some cases, clinical panels may exclude genes with high evidence of disease causality, potentially resulting in failure to identify a patient’s true genetic cause. In other cases, there are genes included in clinical panels with minimal evidence of being truly associated with ALS, potentially resulting in many variants of uncertain significance, defined as having unknown clinical consequences. Variants of uncertain significance may cause unnecessary stress for patients and their families, drain laboratory and clinician time, complicate genetic counselling, and result in patients being excluded from participating in a drug trial due to variant-based exclusion criteria (10, 15). Wide genetic screening in a clinical setting that is not supported by rigorous scientific evidence for causality can hence result in serious consequences and cost valuable resources for laboratories.

In 2017, the NIH-funded Clinical Genome Resource (ClinGen) published a standard framework for the evaluation of gene-disease relationships, and efforts are currently ongoing to classify gene relationships across many diseases by Gene Curation Expert Panels (GCEPs). Using genetic and experimental evidence from the literature, a framework is used to assign scores to genes that have previously been reported to be associated with the diseases, which further correspond to qualitative classification levels for each gene-disease relationship (16). The Amyotrophic Lateral Sclerosis Spectrum Disorders GCEP was formed in 2021 to evaluate the strength of the evidence for genes that have been previously asserted to cause ALS with the central goal of providing evidence based guidance for the standardization of clinical genetic testing worldwide, and therefore, diagnostics and clinical care of ALS patients. As a part of this effort, here we assess the current landscape of the clinical genetic testing panels offered globally and consider the necessity of standardizing classification of gene-disease relationships in ALS.

## Methods

### ALS Clinical Genetic Testing Panel Query

We first interrogated the National Institutes of Health (NIH) Genetic Testing Registry (GTR) (February 2022) to identify the commercial clinical genetic testing panels currently offered globally (17). The search string “amyotrophic lateral sclerosis” was used to set the condition, for which all “Clinical tests” were examined. The “CLIA Certified” filter was applied to select for only those clinical panels that have been Clinical Laboratory Improvement Amendments (CLIA) certified, which is a regulation for laboratory testing issued by the Center for Medicare and Medicaid Services. Members of the ClinGen ALS GCEP were asked to identify any additional commercial genetic panels used by their clinics to assess the genetic causes of ALS patients that had not been captured by the NIH GTR or US-based CLIA certification.

We selected panels specific to “ALS” or “motor neuron disease” and excluded those that spanned other phenotypes (e.g., neurodegenerative disease panels, neuromuscular disease panels). If a company offered multiple panels for ALS, the most comprehensive panel was selected for further analysis. Data were collected on all selected clinical panels, including company location, methods included in the testing, and gene coverage. All panels were reviewed to determine inclusion or exclusion of the *C9orf72* GGGGCC HRE.

### Assessment of Genes Included on Clinical Testing Panels

We further assessed all genes included in the identified clinical genetic panels to determine the encoded protein’s function and whether the genes had previously been associated directly with ALS in the published literature. More specifically, PubMed and Google Scholar were reviewed by searching each gene name in a preliminary search with “ALS”, followed by a secondary search with “motor neuron disease” (March 2022). For a gene to have been considered previously associated with ALS, at least one peer-reviewed study must have identified a variant in the gene in an ALS patient or must have reported a significant association between the gene and phenotype in a case-control analysis. If they indeed had been previously associated with ALS, we examined when and with which study design the first reported relationship was made. We also prioritized the genes to identify those more probably associated with ALS, defined as genes with ≥2 publications asserting rare variants as causative of ALS.

### Assessment of Variants in ALS Genes

We reviewed the NIH ClinVar database for all genes included in the analyzed clinical testing panels and for the identified genes that were not included in any clinical genetic panels but were recently associated with ALS using large-scale association studies (18, 19). The ClinVar variant summary was downloaded from the file transfer protocol site on May 4, 2022 (https://ftp.ncbi.nlm.nih.gov/pub/clinvar/). We identified the number of variants reported in by ClinVar these genes as associated to either the “ALS” or “motor neuron disease” phenotype with one of the following classifications: pathogenic, likely pathogenic, risk factor, benign, likely benign, conflicting interpretation of pathogenicity, other, or uncertain significance. Similarly, variant pathogenicity data was obtained from a commercial genetic testing company (June 2020), and the pathogenicity classification of variants from each gene included in the panel were assessed.

## Results

In total, we identified 33 unique clinical genetic testing panels that encompassed the ALS phenotype, each offered by one of 16 commercial genetic testing companies. Of those, 14 clinical panels were specific to “ALS” or “motor neuron disease” (Table 1). Of the 14 panels, only seven also included, or had the option of including, *C9orf72* GGGGCC HRE analysis. Nearly all panels were offered by commercial companies located in Europe or North America (specifically, the Unites States), although a single test was offered by a company from South Korea.

**Table 1.** Commercial clinical genetic tests offered globally specific to amyotrophic lateral sclerosis (ALS) and motor neuron disease

| **Company** | **Location** | **Test Name** | **GTR Test ID** | **Methods Included** | **# Genes Covered** | ***C9orf72* Expansion** |
| --- | --- | --- | --- | --- | --- | --- |
| Blueprint Genetics | Espoo, Southern Finland, Finland | Amyotrophic lateral sclerosis panel | GTR000552738.3 | NGS of coding region; deletion/duplication | 32 | No |
| CeGaT GmbH | Tübingen, Baden-Württemberg, Germany | Amyotrophic lateral sclerosis panel | GTR000522444.2 | NGS of coding region | 54 | Optional |
| Centogene AG - the Rare Disease Company | Rostock, Mecklenburg-Vorpommern, Germany | Amyotrophic lateral sclerosis panel | GTR000503152.2 | NGS of coding region; deletion/duplication | 22 | Yes |
| Connective Tissue Gene Tests | Allentown, Pennsylvania, USA | Amyotrophic lateral sclerosis and related disorders comprehensive panel | GTR000592010.1 | NGS of coding region; deletion/duplication | 24 | No |
| FirmaLab | Los Angeles, California, USA | Amyotrophic lateral sclerosis sequencing panel | GTR000508862.3 | NGS of coding region | 4 | No |
| Fulgent Genetics | Temple City, California, USA | Amyotrophic lateral sclerosis NGS panel | GTR000509393.12 | NGS of coding region; deletion/duplication | 43 | Yes |
| GC Genome | South Korea | Amyotrophic lateral sclerosis panel | NA | NGS of coding region; deletion/duplication | 27 | No |
| GeneDx | Gaithersburg, Maryland, USA | Amyotrophic lateral sclerosis/frontotemporal lobar degeneration panel | GTR000569718.1 | NGS of coding region; deletion/duplication | 24 | No |
| Invitae | San Francisco, California, USA | Amyotrophic lateral sclerosis Panel with *C9orf72* | GTR000553755.2 | NGS of coding region; deletion/duplication | 22 | Yes |
| Knight Diagnostic Laboratories - Molecular Diagnostic Center | Portland, Oregon, USA | Amyotrophic lateral sclerosis | GTR000552113.1 | NGS of coding region | 21 | No |
| Mayo Clinical Laboratories | Rochester, Minnesota, USA | Motor neuron disease panel | GTR000568254.3 | NGS of coding region | 17 | No |
| Medical Neurogenetics Laboratories, LLC. | Atlanta, Georgia, USA | Amyotrophic lateral sclerosis NGS panel | GTR000559549.2 | NGS of coding region | 30 | No |
| PreventionGenetics | Marshfield, Wisconsin, USA | Amyotrophic lateral sclerosis panel | GTR000591896.1 | NGS of coding region; deletion/duplication | 33 | Yes |
| SciLifeUAS | Uppsala, Sweden | SciLifeUppsala-Um | NA | NGS of coding region; enzymatic assay in all *SOD1* cases | 49 | Yes |

Only tests specific to the ALS or motor neuron disease phenotype were included, although panels relevant for both ALS and frontotemporal dementia were retained FTD due to the large amount of genetic overlap and clinical co-occurrence between the two phenotypes. In selecting clinical panels for review, if multiple panels were offered by the company for genetic testing of ALS, the most comprehensive test was chosen. Abbreviations: GTR, Genetic Testing Registry; NA, not applicable; NGS, next-generation sequencing; #, number; USA, United States of America.

Table 2 compares the 91 different genes covered by the ALS specific commercial clinical genetic testing panels. The commonly included were *ANG, FUS, OPTN, SETX, SOD1, TARDBP, UBQLN2, VAPB*, and *VCP* (≥13 tests). By contrast, 40 genes were found on a single panel, and 14 genes had not previously been directly associated with “ALS” or “motor neuron disease” in the literature (Table 3). We also identified 44 genes with ≥2 publications reporting rare coding variants as associated with ALS (Figure 1). The additional reports replicating the original findings may suggest a greater probability of the genes having a true relationship with ALS; however, do not definitively define causality. Figure 2 shows the relationship between year of discovery and the number of panels each gene is included on.

**Table 2.**
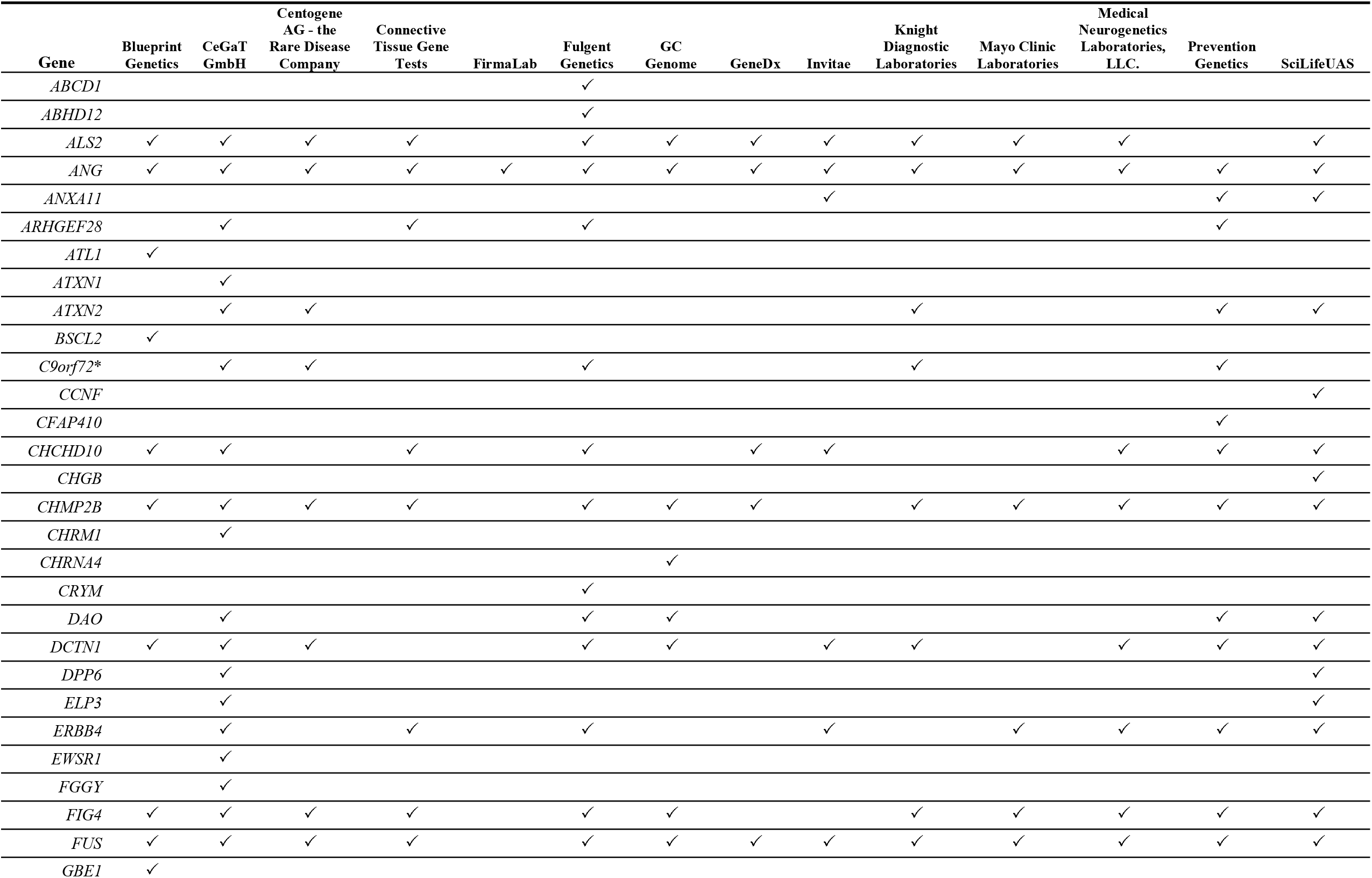
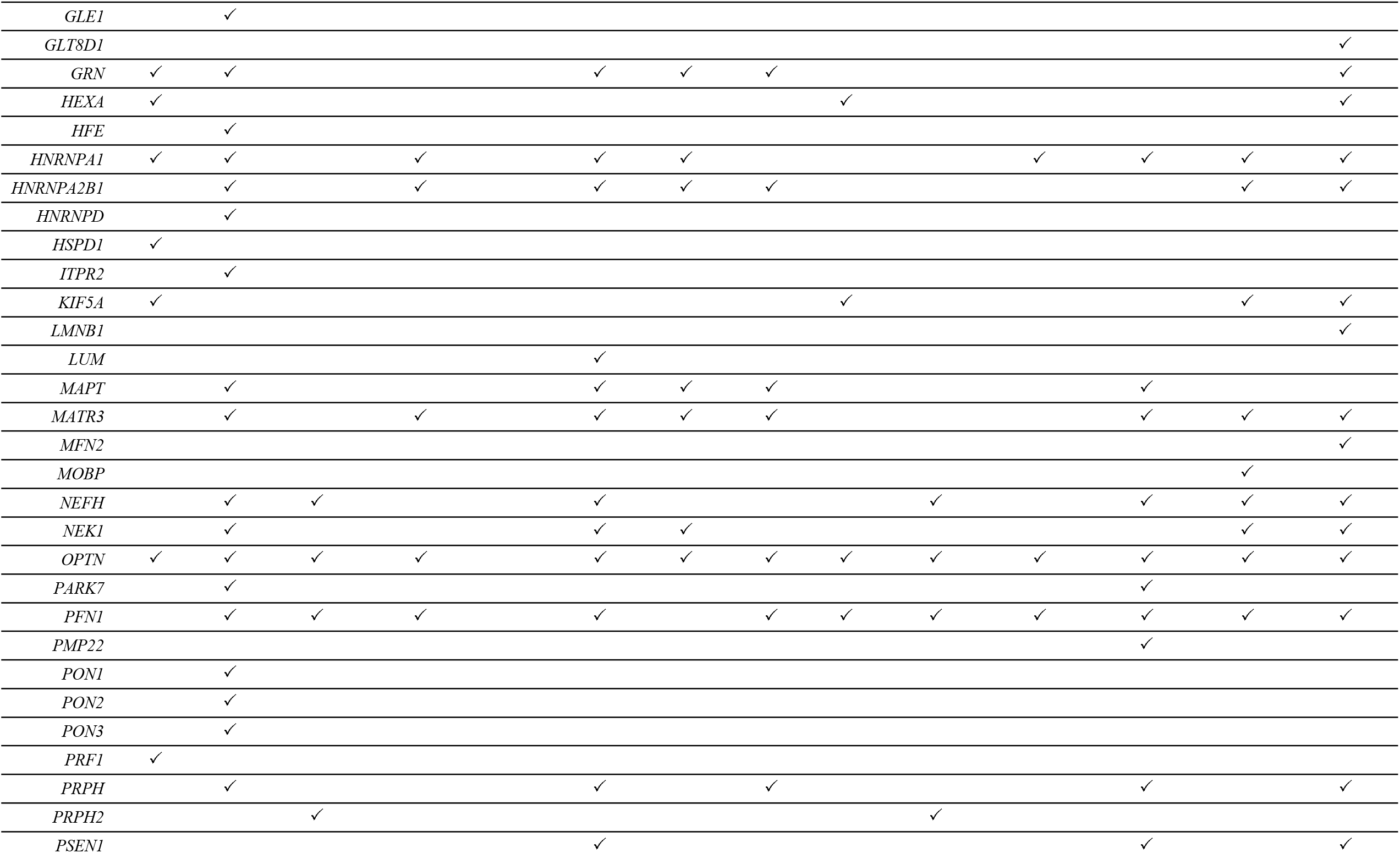
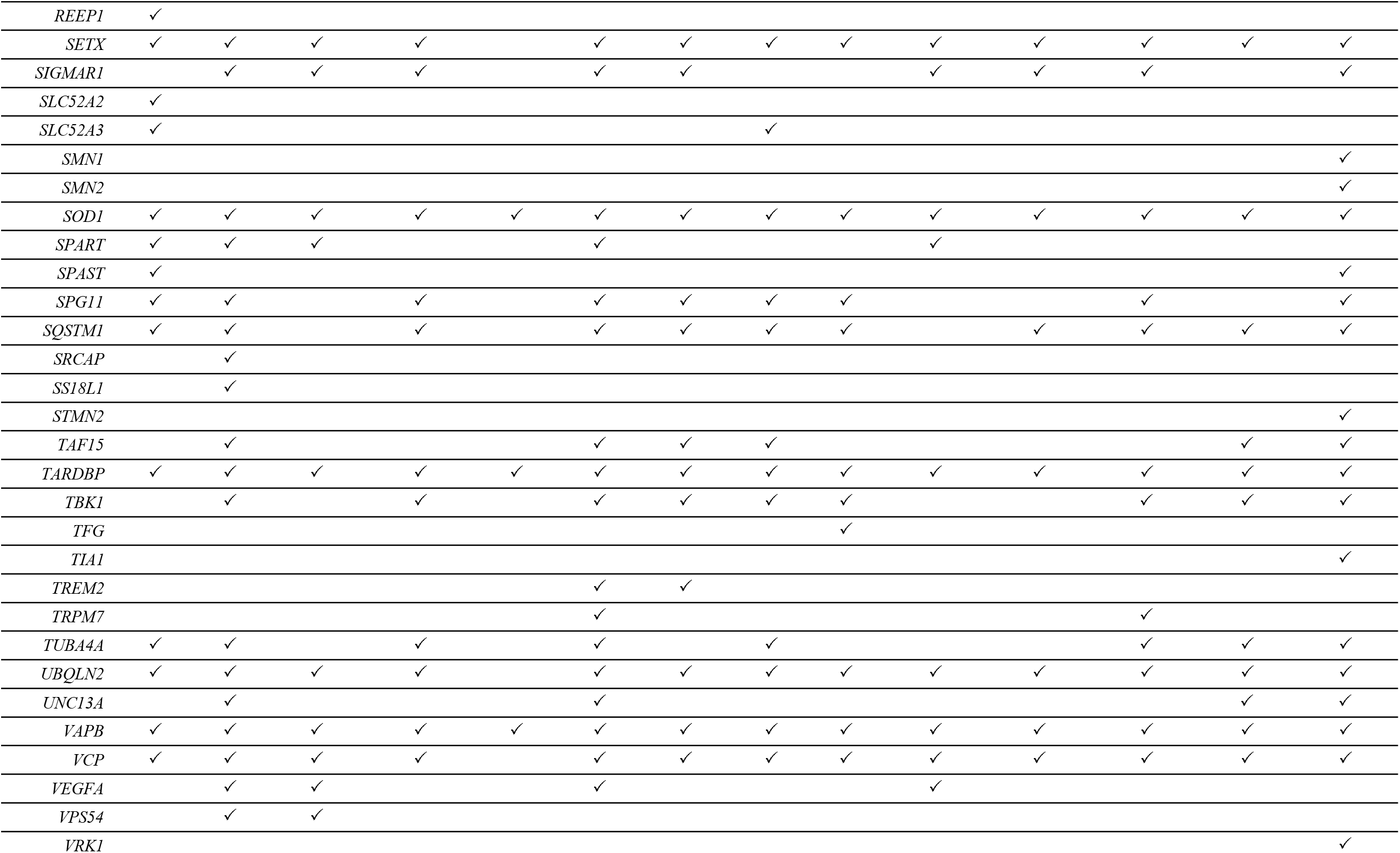
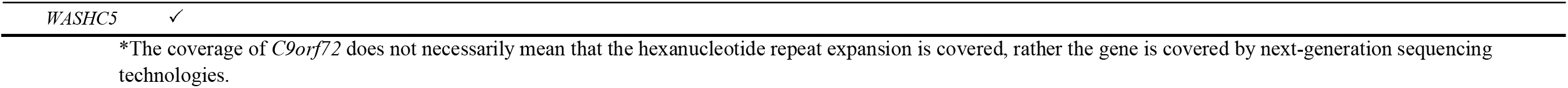
Genes covered by the commercial clinical genetic tests offered globally specific to amyotrophic lateral sclerosis (ALS)

| Gene | Blueprint Genetics | CeGaT GmbH | Centogene AG - the Rare Disease Company | Connective Tissue Gene Tests | FirmaLab | Fulgent Genetics | GC Genome | GeneDx | Invitae | Knight Diagnostic Laboratories | Mayo Clinic Laboratories | Medical Neurogenetics Laboratories, LLC. | Prevention Genetics | SciLifeUAS |
| --- | --- | --- | --- | --- | --- | --- | --- | --- | --- | --- | --- | --- | --- | --- |
| *ABCD1* |  |  |  |  |  | ✓ |  |  |  |  |  |  |  |  |
| *ABHD12* |  |  |  |  |  | ✓ |  |  |  |  |  |  |  |  |
| *ALS2* | ✓ | ✓ | ✓ | ✓ |  | ✓ | ✓ | ✓ | ✓ | ✓ | ✓ | ✓ |  | ✓ |
| *ANG* | ✓ | ✓ | ✓ | ✓ | ✓ | ✓ | ✓ | ✓ | ✓ | ✓ | ✓ | ✓ | ✓ | ✓ |
| *ANXA11* |  |  |  |  |  |  |  |  | ✓ |  |  |  | ✓ | ✓ |
| *ARHGEF28* |  | ✓ |  | ✓ |  | ✓ |  |  |  |  |  |  | ✓ |  |
| *ATL1* | ✓ |  |  |  |  |  |  |  |  |  |  |  |  |  |
| *ATXN1* |  | ✓ |  |  |  |  |  |  |  |  |  |  |  |  |
| *ATXN2* |  | ✓ | ✓ |  |  |  |  |  |  | ✓ |  |  | ✓ | ✓ |
| *BSCL2* | ✓ |  |  |  |  |  |  |  |  |  |  |  |  |  |
| *C9orf72\** |  | ✓ | ✓ |  |  | ✓ |  |  |  | ✓ |  |  | ✓ |  |
| *CCNF* |  |  |  |  |  |  |  |  |  |  |  |  |  | ✓ |
| *CFAP410* |  |  |  |  |  |  |  |  |  |  |  |  | ✓ |  |
| *CHCHD10* | ✓ | ✓ |  | ✓ |  | ✓ |  | ✓ | ✓ |  |  | ✓ | ✓ | ✓ |
| *CHGB* |  |  |  |  |  |  |  |  |  |  |  |  |  | ✓ |
| *CHMP2B* | ✓ | ✓ | ✓ | ✓ |  | ✓ | ✓ | ✓ |  | ✓ | ✓ | ✓ | ✓ | ✓ |
| *CHRM1* |  | ✓ |  |  |  |  |  |  |  |  |  |  |  |  |
| *CHRNA4* |  |  |  |  |  |  | ✓ |  |  |  |  |  |  |  |
| *CRYM* |  |  |  |  |  | ✓ |  |  |  |  |  |  |  |  |
| *DAO* |  | ✓ |  |  |  | ✓ | ✓ |  |  |  |  |  | ✓ | ✓ |
| *DCTN1* | ✓ | ✓ | ✓ |  |  | ✓ | ✓ |  | ✓ | ✓ |  | ✓ | ✓ | ✓ |
| *DPP6* |  | ✓ |  |  |  |  |  |  |  |  |  |  |  | ✓ |
| *ELP3* |  | ✓ |  |  |  |  |  |  |  |  |  |  |  | ✓ |
| *ERBB4* |  | ✓ |  | ✓ |  | ✓ |  |  | ✓ |  | ✓ | ✓ | ✓ | ✓ |
| *EWSR1* |  | ✓ |  |  |  |  |  |  |  |  |  |  |  |  |
| *FGGY* |  | ✓ |  |  |  |  |  |  |  |  |  |  |  |  |
| *FIG4* | ✓ | ✓ | ✓ | ✓ |  | ✓ | ✓ |  |  | ✓ | ✓ | ✓ | ✓ | ✓ |
| *FUS* | ✓ | ✓ | ✓ | ✓ |  | ✓ | ✓ | ✓ | ✓ | ✓ | ✓ | ✓ | ✓ | ✓ |
| *GBE1* | ✓ |  |  |  |  |  |  |  |  |  |  |  |  |  |
| *GLE1* |  | ✓ |  |  |  |  |  |  |  |  |  |  |  |  |
| *GLT8D1* |  |  |  |  |  |  |  |  |  |  |  |  |  | ✓ |
| *GRN* | ✓ | ✓ |  |  |  | ✓ | ✓ | ✓ |  |  |  |  |  | ✓ |
| *HEXA* | ✓ |  |  |  |  |  |  |  | ✓ |  |  |  |  | ✓ |
| *HFE* |  | ✓ |  |  |  |  |  |  |  |  |  |  |  |  |
| *HNRNPA1* | ✓ | ✓ |  | ✓ |  | ✓ | ✓ |  |  |  | ✓ | ✓ | ✓ | ✓ |
| *HNRNPA2B1* |  | ✓ |  | ✓ |  | ✓ | ✓ | ✓ |  |  |  |  | ✓ | ✓ |
| *HNRNPD* |  | ✓ |  |  |  |  |  |  |  |  |  |  |  |  |
| *HSPD1* | ✓ |  |  |  |  |  |  |  |  |  |  |  |  |  |
| *ITPR2* |  | ✓ |  |  |  |  |  |  |  |  |  |  |  |  |
| *KIF5A* | ✓ |  |  |  |  |  |  |  | ✓ |  |  |  | ✓ | ✓ |
| *LMNB1* |  |  |  |  |  |  |  |  |  |  |  |  |  | ✓ |
| *LUM* |  |  |  |  |  | ✓ |  |  |  |  |  |  |  |  |
| *MAPT* |  | ✓ |  |  |  | ✓ | ✓ | ✓ |  |  |  | ✓ |  |  |
| *MATR3* |  | ✓ |  | ✓ |  | ✓ | ✓ | ✓ |  |  |  | ✓ | ✓ | ✓ |
| *MFN2* |  |  |  |  |  |  |  |  |  |  |  |  |  | ✓ |
| *MOBP* |  |  |  |  |  |  |  |  |  |  |  |  | ✓ |  |
| *NEFH* |  | ✓ | ✓ |  |  | ✓ |  |  |  | ✓ |  | ✓ | ✓ | ✓ |
| *NEK1* |  | ✓ |  |  |  | ✓ | ✓ |  |  |  |  |  | ✓ | ✓ |
| *OPTN* | ✓ | ✓ | ✓ | ✓ |  | ✓ | ✓ | ✓ | ✓ | ✓ | ✓ | ✓ | ✓ | ✓ |
| *PARK7* |  | ✓ |  |  |  |  |  |  |  |  |  | ✓ |  |  |
| *PFN1* |  | ✓ | ✓ | ✓ |  | ✓ |  | ✓ | ✓ | ✓ | ✓ | ✓ | ✓ | ✓ |
| *PMP22* |  |  |  |  |  |  |  |  |  |  |  | ✓ |  |  |
| *PON1* |  | ✓ |  |  |  |  |  |  |  |  |  |  |  |  |
| *PON2* |  | ✓ |  |  |  |  |  |  |  |  |  |  |  |  |
| *PON3* |  | ✓ |  |  |  |  |  |  |  |  |  |  |  |  |
| *PRF1* | ✓ |  |  |  |  |  |  |  |  |  |  |  |  |  |
| *PRPH* |  | ✓ |  |  |  | ✓ |  | ✓ |  |  |  | ✓ |  | ✓ |
| *PRPH2* |  |  | ✓ |  |  |  |  |  |  | ✓ |  |  |  |  |
| *PSEN1* |  |  |  |  |  | ✓ |  |  |  |  |  | ✓ |  | ✓ |
| *REEP1* | ✓ |  |  |  |  |  |  |  |  |  |  |  |  |  |
| *SETX* | ✓ | ✓ | ✓ | ✓ |  | ✓ | ✓ | ✓ | ✓ | ✓ | ✓ | ✓ | ✓ | ✓ |
| *SIGMAR1* |  | ✓ | ✓ | ✓ |  | ✓ | ✓ |  |  | ✓ | ✓ | ✓ |  | ✓ |
| *SLC52A2* | ✓ |  |  |  |  |  |  |  |  |  |  |  |  |  |
| *SLC52A3* | ✓ |  |  |  |  |  |  | ✓ |  |  |  |  |  |  |
| *SMN1* |  |  |  |  |  |  |  |  |  |  |  |  |  | ✓ |
| *SMN2* |  |  |  |  |  |  |  |  |  |  |  |  |  | ✓ |
| *SOD1* | ✓ | ✓ | ✓ | ✓ | ✓ | ✓ | ✓ | ✓ | ✓ | ✓ | ✓ | ✓ | ✓ | ✓ |
| *SPART* | ✓ | ✓ | ✓ |  |  | ✓ |  |  |  | ✓ |  |  |  |  |
| *SPAST* | ✓ |  |  |  |  |  |  |  |  |  |  |  |  | ✓ |
| *SPG11* | ✓ | ✓ |  | ✓ |  | ✓ | ✓ | ✓ | ✓ |  |  | ✓ |  | ✓ |
| *SQSTM1* | ✓ | ✓ |  | ✓ |  | ✓ | ✓ | ✓ | ✓ |  | ✓ | ✓ | ✓ | ✓ |
| *SRCAP* |  | ✓ |  |  |  |  |  |  |  |  |  |  |  |  |
| *SS18L1* |  | ✓ |  |  |  |  |  |  |  |  |  |  |  |  |
| *STMN2* |  |  |  |  |  |  |  |  |  |  |  |  |  | ✓ |
| *TAF15* |  | ✓ |  |  |  | ✓ | ✓ | ✓ |  |  |  |  | ✓ | ✓ |
| *TARDBP* | ✓ | ✓ | ✓ | ✓ | ✓ | ✓ | ✓ | ✓ | ✓ | ✓ | ✓ | ✓ | ✓ | ✓ |
| *TBK1* |  | ✓ |  | ✓ |  | ✓ | ✓ | ✓ | ✓ |  |  | ✓ | ✓ | ✓ |
| *TFG* |  |  |  |  |  |  |  |  | ✓ |  |  |  |  |  |
| *TIA1* |  |  |  |  |  |  |  |  |  |  |  |  |  | ✓ |
| *TREM2* |  |  |  |  |  | ✓ | ✓ |  |  |  |  |  |  |  |
| *TRPM7* |  |  |  |  |  | ✓ |  |  |  |  |  | ✓ |  |  |
| *TUBA4A* | ✓ | ✓ |  | ✓ |  | ✓ |  | ✓ |  |  |  | ✓ | ✓ | ✓ |
| *UBQLN2* | ✓ | ✓ | ✓ | ✓ |  | ✓ | ✓ | ✓ | ✓ | ✓ | ✓ | ✓ | ✓ | ✓ |
| *UNC13A* |  | ✓ |  |  |  | ✓ |  |  |  |  |  |  | ✓ | ✓ |
| *VAPB* | ✓ | ✓ | ✓ | ✓ | ✓ | ✓ | ✓ | ✓ | ✓ | ✓ | ✓ | ✓ | ✓ | ✓ |
| *VCP* | ✓ | ✓ | ✓ | ✓ |  | ✓ | ✓ | ✓ | ✓ | ✓ | ✓ | ✓ | ✓ | ✓ |
| *VEGFA* |  | ✓ | ✓ |  |  | ✓ |  |  |  | ✓ |  |  |  |  |
| *VPS54* |  | ✓ | ✓ |  |  |  |  |  |  |  |  |  |  |  |
| *VRK1* |  |  |  |  |  |  |  |  |  |  |  |  |  | ✓ |

| Gene | Blueprint<br>Genetics | CeGaT<br>GmbH | Centogene<br>AG - the<br>Rare Disease<br>Company | Connective<br>Tissue Gene<br>Tests | FirmaLab | Fulgent<br>Genetics | GC<br>Genome | GeneDx | Invitae | Knight<br>Diagnostic<br>Laboratories | Mayo Clinic<br>Laboratories | Medical<br>Neurogenetics<br>Laboratories,<br>LLC. | Prevention<br>Genetics | SciLifeUAS |
| --- | --- | --- | --- | --- | --- | --- | --- | --- | --- | --- | --- | --- | --- | --- |
| *WASHC5* | ✓ |  |  |  |  |  |  |  |  |  |  |  |  |  |

\*The coverage of *C9orf72* does not necessarily mean that the hexanucleotide repeat expansion is covered, rather the gene is covered by next-generation sequencing technologies.

**Table 3.**
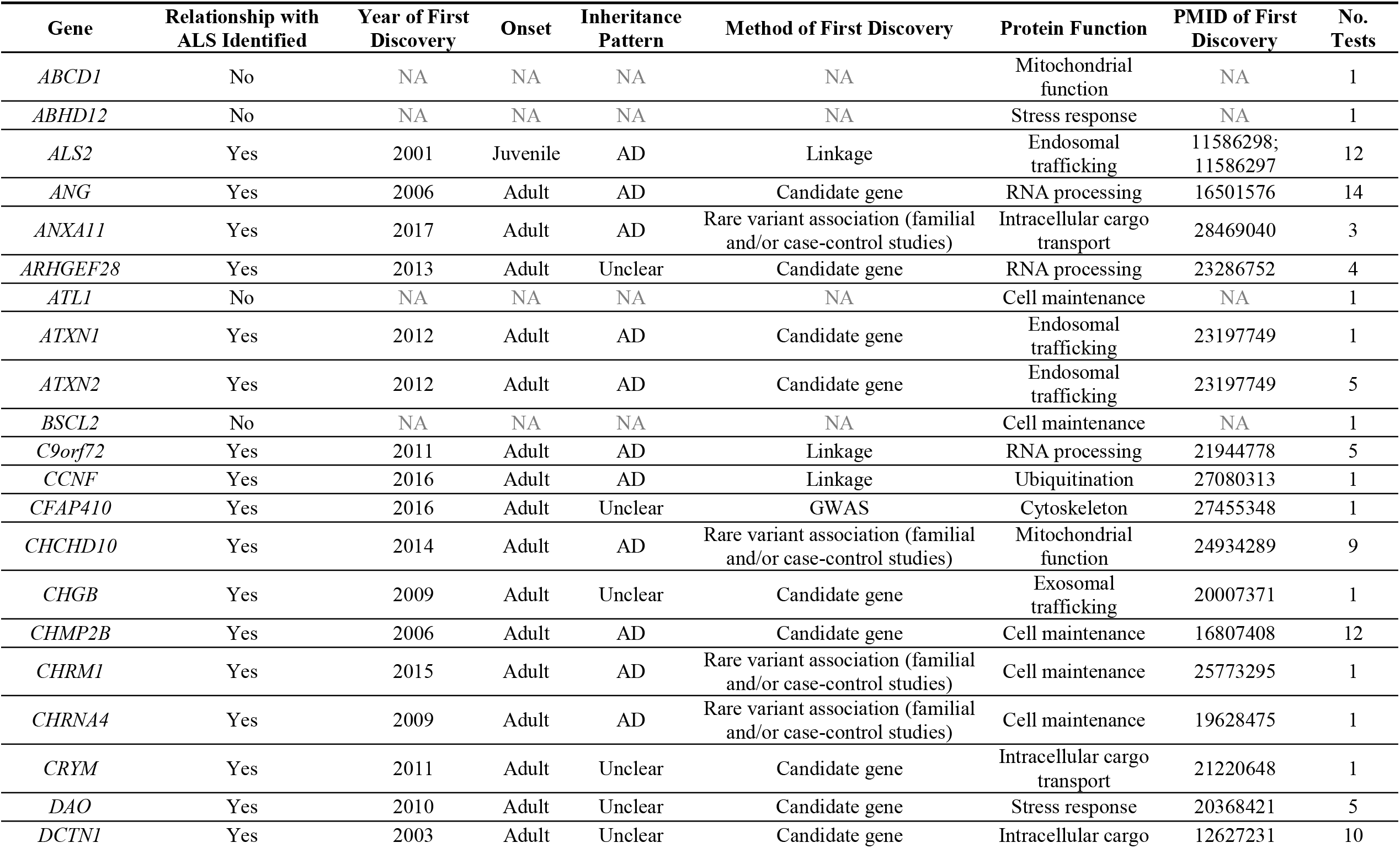
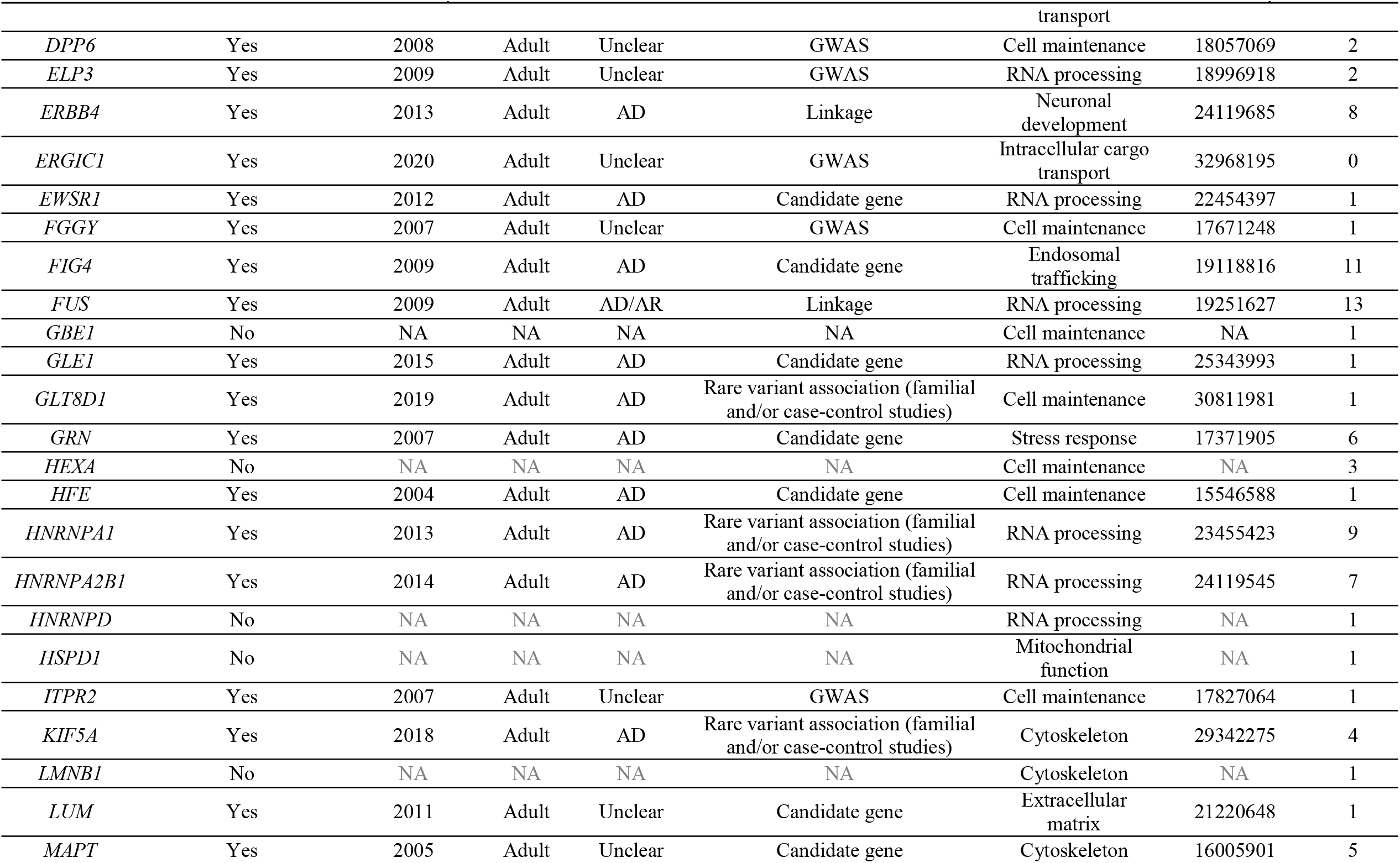
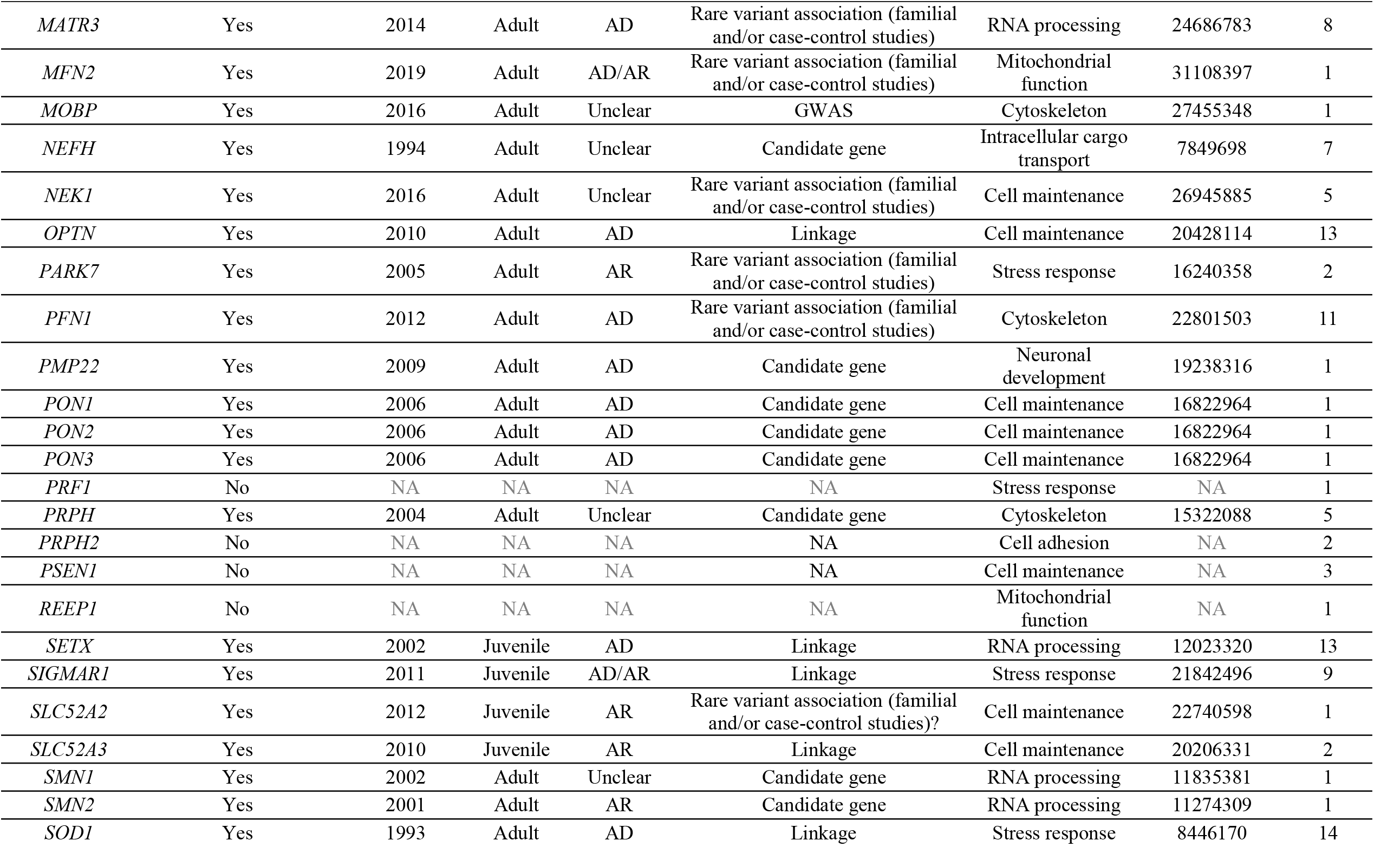
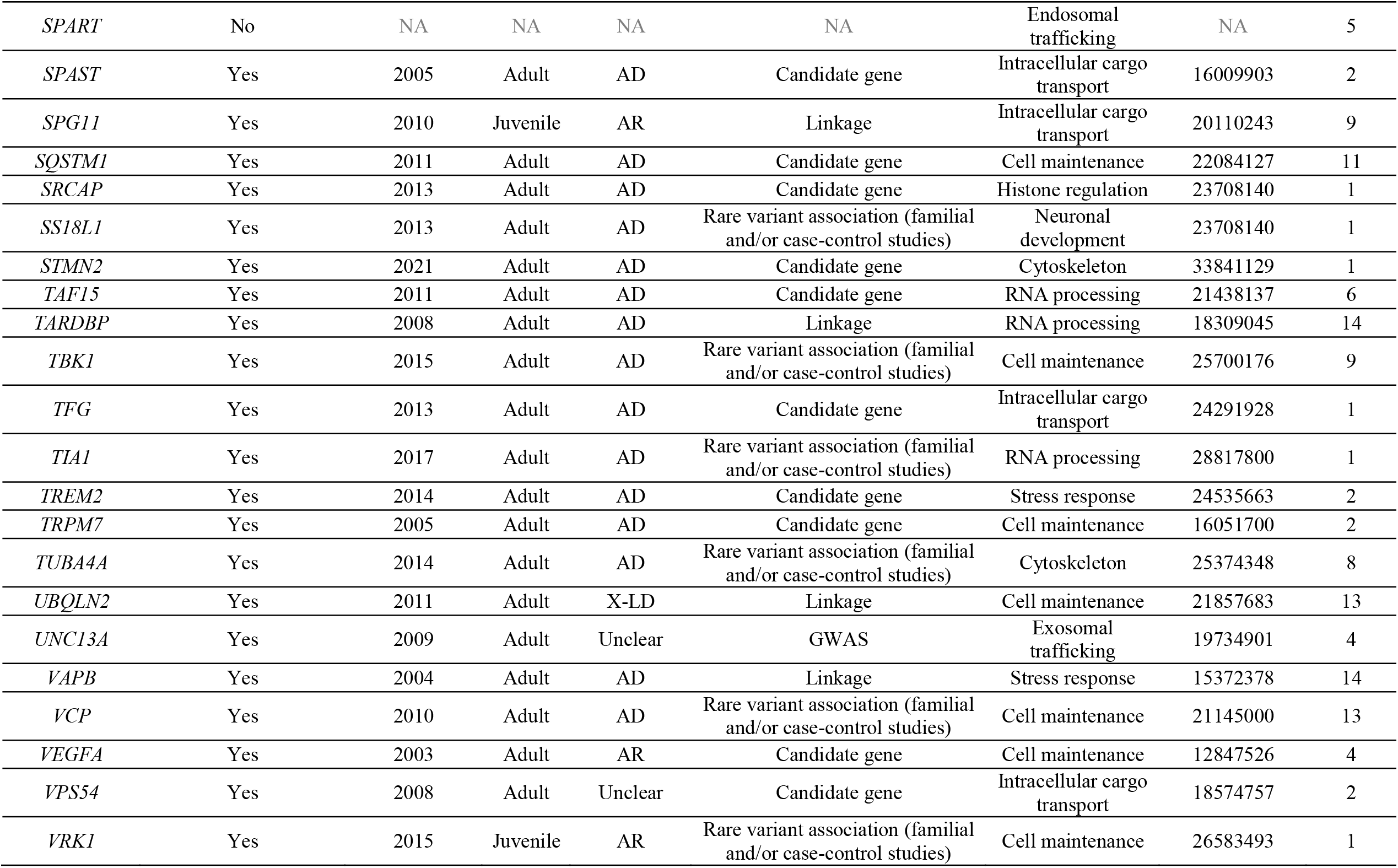
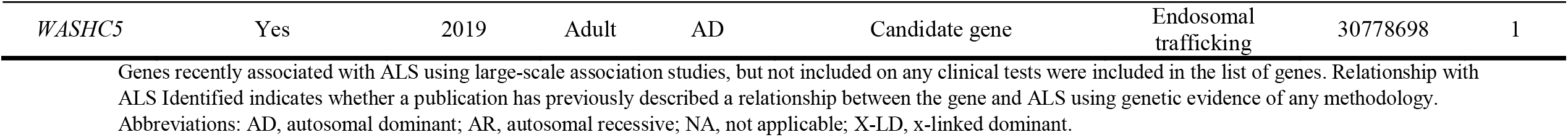
Characteristics of the genes encompassed by the commercial clinical genetic tests offered globally specific to amyotrophic lateral sclerosis (ALS), as well as potential genes of interest missing from clinical genetic tests

**Figure 1.**
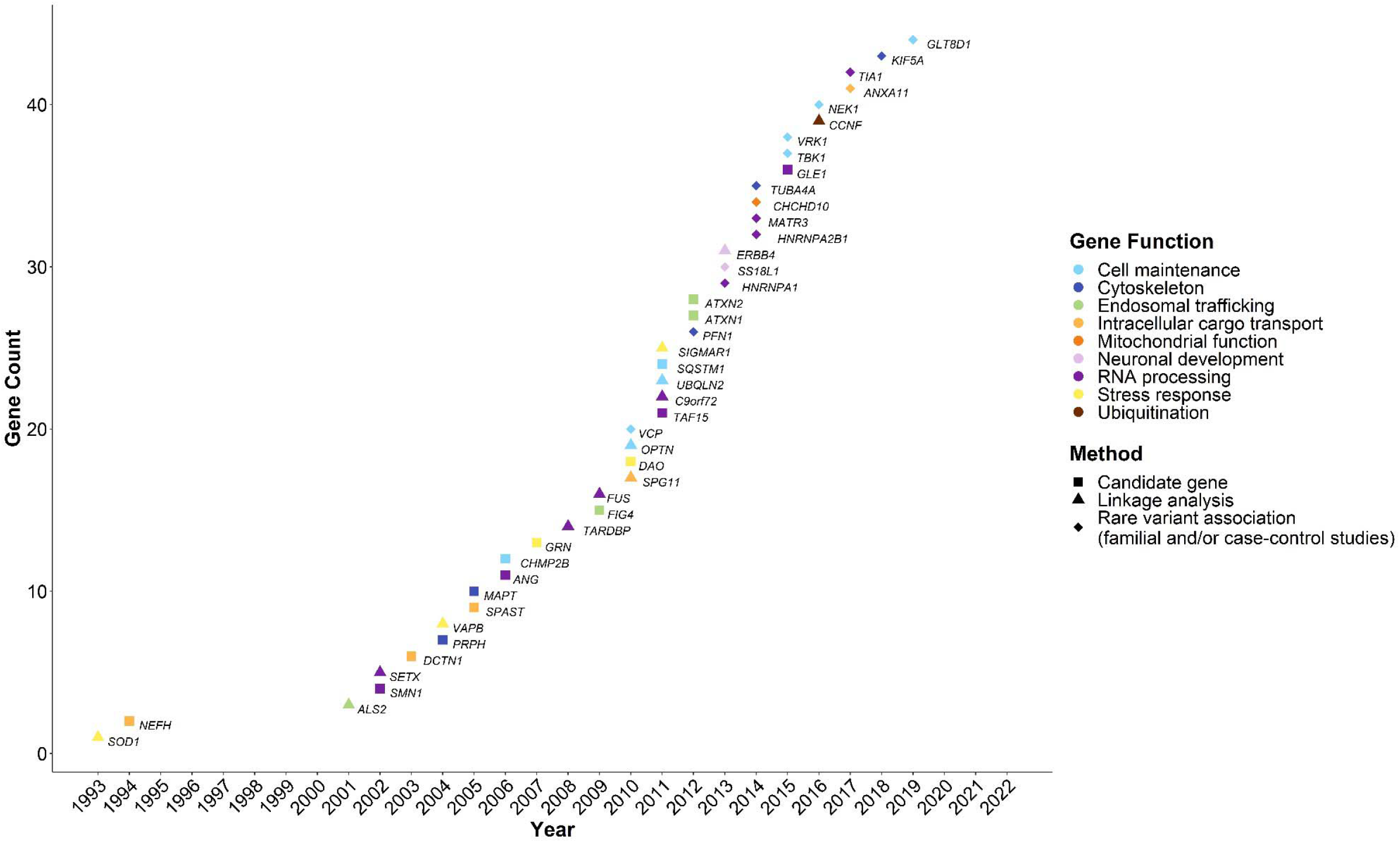
Year of first discovery of a relationship with amyotrophic lateral sclerosis (ALS), method of discovery, and function of the encoded protein of the genes included on the identified ALS specific commercial clinical genetic tests (N=14) with ≥2 publications reporting rare variants associated with ALS.

**Figure 2.**
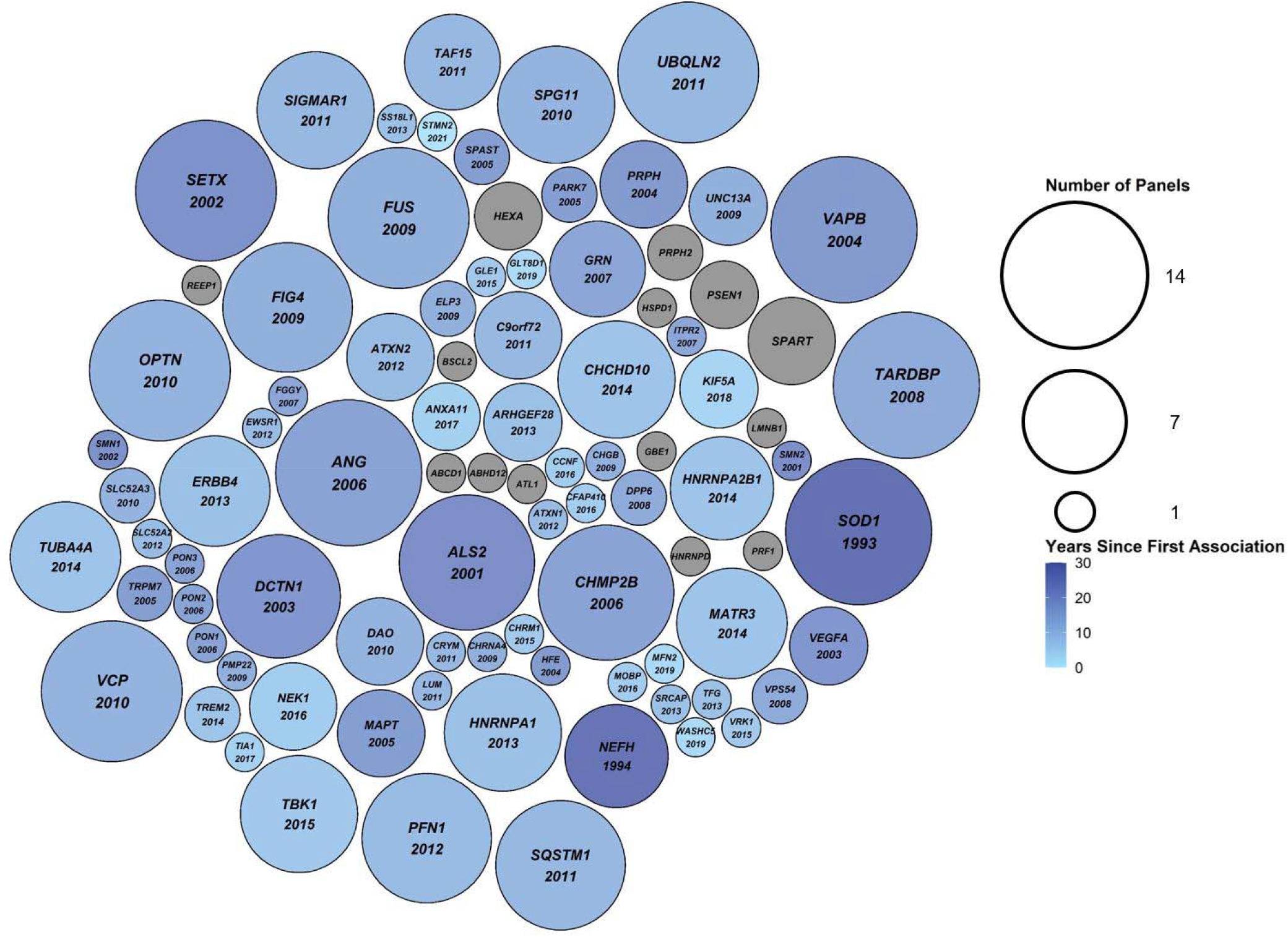
Packed circle plot comparing the number of amyotrophic lateral sclerosis (ALS) specific commercial clinical genetic tests (N=14) each gene was included on. The size of the circles corresponds to the number of clinical genetic tests the gene was included on, ranging from one to 14 tests. The colour of the circles corresponds to the number of years since the gene’s first association with ALS was made. Grey circles indicate genes that have never been associated with ALS. The year of first association between ALS and the gene is indicated below the gene name, where applicable.

Finally, we reviewed ClinVar — a database for clinical and research laboratories to upload how they have classified variants found using genetic testing — to investigate the distribution of variants and their classifications for each of the genes included on any panel. Of the 91 genes, 50 contained variants reported in ClinVar (Figure 3A). *SOD1, TBK1, FUS*, and *OPTN* had the most likely pathogenic or pathogenic variant classifications for ALS (75, 31, 22, and 22 variants, respectively). *SETX, DCTN1*, and *SQSTM1* had the most total variants, (567, 508, and 252 variants, respectively); however, the vast majority were of uncertain significance (403 [71.1%], 364 [71.7%], and 172 [68.3%], respectively). Most variants of uncertain significance in ALS genes were missense variants, including 344 (85.4%) missense variants in *SETX*, 310 (85.2%) missense variants in *DCTN1*, and 154 (89.5%) missense variants in *SQSTM1* (Figure 3C). Across all variants of uncertain significance (n = 2095), only ∼3% (n = 63) were considered protein truncating variants (including frameshift insertions/deletions, splicing variants, and stop gain variants), whereas across all likely pathogenic/pathogenic variants (n = 256) over 28% (n = 72) were considered protein truncating variants (Figure 3B). A similar exercise was performed using data from a commercial genetic testing company (Figure 4). Here, *SETX, ARHGEF28*, and *SQSTM1* had the most variants of any classification, with 137, 90, and 80 variants, respectively, yet in contrast to the ClinVar data, most variants in these genes were considered likely benign (82 [59.9%], 68 [75.6%], and 42 [52.5%], respectively).

**Figure 3.**
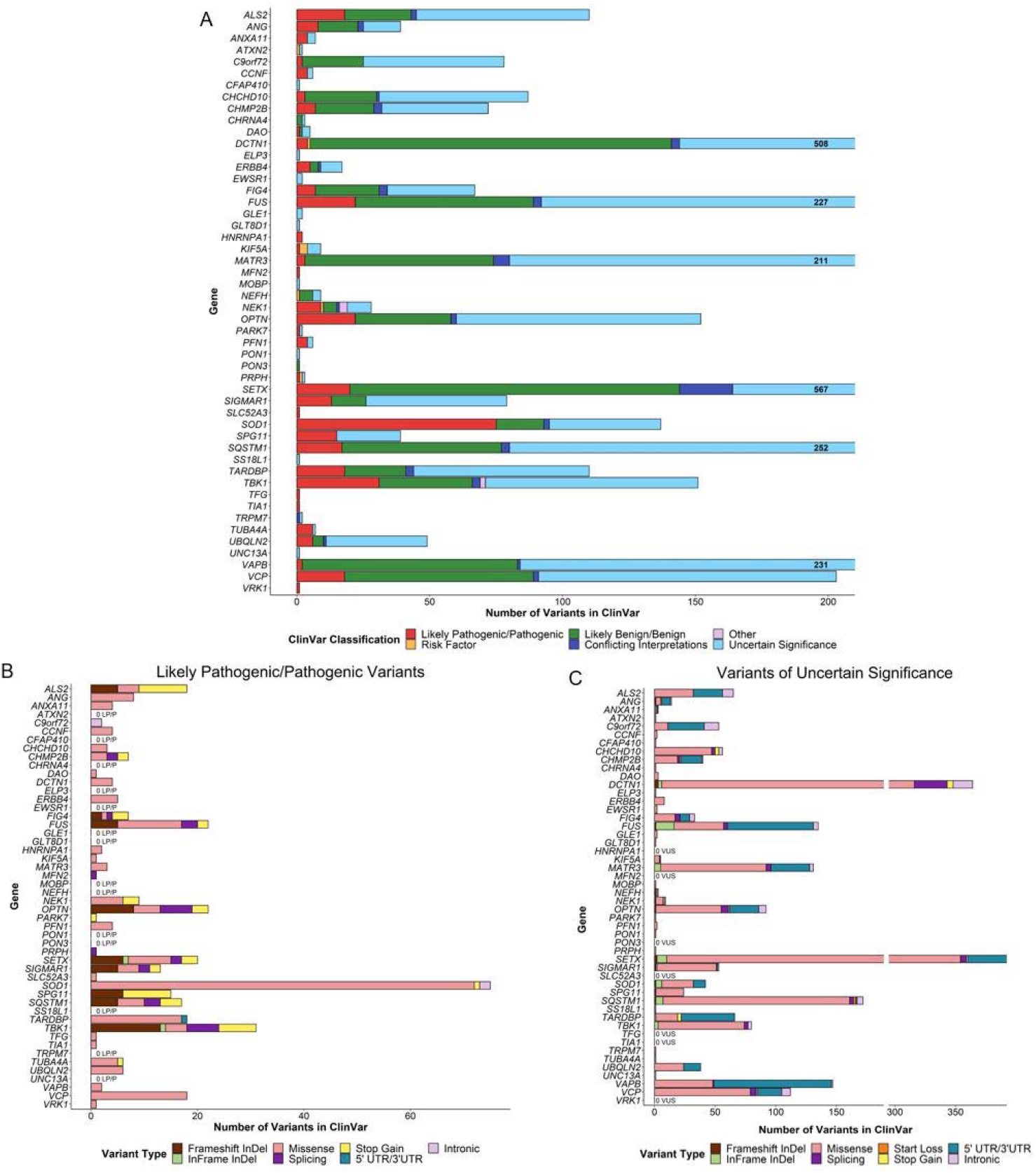
Pathogenicity classification and variant types reported in ClinVar in genes previously associated with amyotrophic lateral sclerosis (ALS). All genes found on at least one of the analyzed clinical genetic testing panels (N=14) were included in the analysis. However, only genes with variants reported as associated with “ALS” or “motor neuron disease” in ClinVar are displayed in the figure. **(A)** Pathogenicity classification of all variants reported in ClinVar as associated with ALS. The total number of variants reported in ClinVar in *DCTN1, FUS, MATR3, SETX, SQSTM1*, and *VAPB* exceeded the y-axis limits of 200; for these genes, the total number of variants found in ClinVar were included on the right side of the bar plot. **(B)** Variant types of all variants reported in ClinVar as likely pathogenic or pathogenic for ALS. **(C)** Variant types of all variants reported in ClinVar as being of uncertain significance for ALS. Abbreviations: LP, likely pathogenic; P, pathogenic; VUS, variants of uncertain significance.

**Figure 4.**
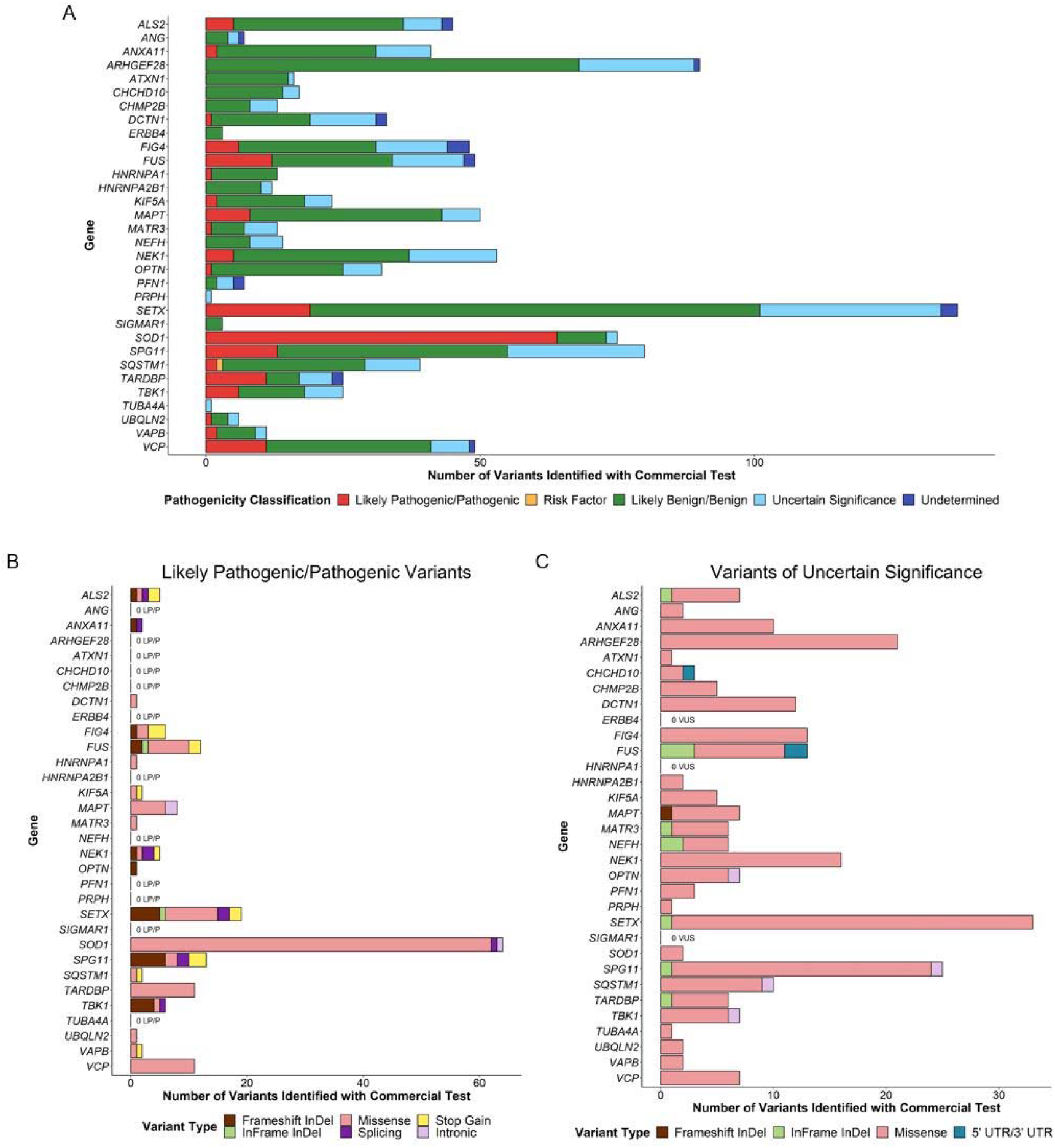
Pathogenicity classification and variant types identified using a commercially available genetic test for amyotrophic lateral sclerosis (ALS). **(A)** Pathogenicity classification of variants identified using a commercially available genetic test for ALS. **(B)** Variant types of all variants identified using a commercially available genetic test for ALS and classified as likely pathogenic or pathogenic. **(C)** Variant types of all variants identified using a commercially available genetic test for ALS and classified as being of uncertain significance. Abbreviations: LP, likely pathogenic; P, pathogenic; VUS, variants of uncertain significance.

## Discussion

The range of clinical genetic testing panels offered globally for the assessment of ALS risk or causality is highly heterogeneous. Ideally, clinical laboratories would use a standardized approach of including genes with greater levels of evidence of gene-disease relationships, resulting in consistency across companies. However, that approach is not reflected in the current landscape. It is the aim of the ClinGen ALS GCEP to provide the necessary resources to move towards the offering of consistent panels world-wide that include genes with high levels of evidence for true gene-disease relationships.

Our analysis shows that a group of core genes are represented on most panels — over 90% of the clinical panels included sequencing of the genes *ANG, FUS, OPTN, SETX, SOD1, TARDBP, UBQLN2, VAPB*, and *VCP*. Further, a recent assessment of Canadian clinics offering genetic testing for fALS cases found that a substantial subset (36%) are only ordering tests for *SOD1* and the *C9orf72* GGGGCC HRE (8), likely resulting in a large number of false negative genetic results from the failure to assess other genes with a definitive relationship with ALS.

Our analysis also shows that many panels are missing genes with large and growing bodies of evidence for their causality in ALS. Examples include *NEK1* and *ANXA11*, each included in <40% of clinical panels regardless of the relatively large amount of genetic and experimental evidence supporting their implication in ALS (20-25). Interestingly, both genes were first reported as associated with ALS within the past six years, fitting the common trend we observed that likelihood of being included on panels seemingly correlated closely with time since discovery. Further, many laboratories have been inconsistent and idiosyncratic in deciding which genes to report on, of the 91 genes included in at least one of the ALS clinical genetic panels analyzed, 40 of the genes (44.0%) were each included in only a single clinical panel, and 14 genes (15.4%) had not been previously directly associated with ALS in the literature. Although a subset of these genes may be associated with other motor neuron disorders, such as hereditary spastic paraplegia, the inclusion of genes in clinical panels lacking strong evidence of a true gene-disease relationship is of particular concern due to the high rates of variants of uncertain significance that can be identified (11) and the confusion that can be cause when genes are reported without a strong phenotypic match. In fact, the genes with the highest rates of variants of uncertain significance in ClinVar — namely, *SETX, DCTN1*, and *SQSTM1* — are each included in at least 10 of the 14 analyzed clinical panels (71.4%). As aforementioned, variants of uncertain significance complicate the clinical testing process due to the inability to draw clinically relevant conclusions from their identification (10, 15).

Only 50% of analyzed panels included, or offered the option of including, *C9orf72* repeat expansion analysis despite being one of the most commonly inherited forms of ALS, including cases of sALS (2-4). While only a subset of the clinical panels included the addition of *C9orf72* HRE screening, the assessment may be ordered separately from a panel, potentially using a tiered approach by ruling out the repeat as a primary cause prior to further testing. Regardless, its inclusion in all clinical testing for ALS is imperative; without it, the genetic diagnosis of a patient may be overlooked in nearly 15% of ALS cases, although rates differ by population.

Panel testing may not be flexible if targeted capture is the primary the method. Evidence that this is the cases comes from the fact that the likelihood of a gene being included on a panel correlates closely with time from discovery; in many instances, genes first associated with ALS longer ago were included in a greater number of panels. In fact, all nine aforementioned genes included in more than 90% of clinical panels were first associated with ALS over 10 years ago. Similarly, the clinical panels do not include potentially relevant genes with more recently identified relationships to ALS, as previously discussed regarding *NEK1* and *ANXA11*. Time since discovery will also influence on the amount of genetic and experiment evidence supporting a gene-disease relationship available to the ALS GCEP for the purposes of gene curation. To account for this, following the initial round of gene curation, monitoring of gene-disease relationships will continue as new genetic and experimental evidence becomes available, and new gene disease relationships are discovered.

It is recognized that the assessment of clinical tests presented herein is limited to only 14 panels, mostly from Europe and North America. Yet, we anticipate that similar heterogeneity would be observed, or even amplified, following the inclusion of tests from underrepresented locations, and that the need for consistency across clinical genetic testing in ALS would remain apparent.

In addition to genes that have previously had a relationship to ALS defined, the ClinGen ALS spectrum disorders GCEP is curating the gene-disease relationships for all genes associated with ALS-spectrum phenotypes, such as progressive muscular atrophy (PMA), primary lateral sclerosis (PLS), and progressive bulbar palsy, as well as genes associated with FTD. Although a future publication will describe the complete curation methodology and overview of the gene-disease relationships defined, an up to date compendium of all gene-disease validity classifications approved by the ALS GCEP is available on the <u>ClinGen Gene-Disease Validity Portal</u>. It is anticipated that the consensus regarding gene-disease relationships will improve the application of genetic testing clinically and, ultimately, the care of patients living with ALS and their families.

## Data Availability

All data produced in the present work are contained in the manuscript.

## Acknowledgements

ClinGen is primarily funded by the National Human Genome Research Institute (NHGRI) with co-funding from the National Cancer Institute (NCI), through the following grants: Baylor/Stanford - U24HG009649, Broad/Geisinger - U24HG006834, and UNC/Kaiser - U24HG009650. The content is solely the responsibility of the authors and does not necessarily represent the official views of the National Institutes of Health. A.A.D. is supported by the Canadian Institute of Health Research Banting Postdoctoral Fellowship program. H.M is supported by GlaxoSmithKline and the KCL funded centre for Doctoral Training (CDT) in Data-Driven Health. I.B. is supported by the National Health and Medical Research Council of Australia (1176913). W.v.R. is supported by funding provided by the Dutch Research Council (NWO) [VENI scheme grant 09150161810018] and Prinses Beatrix Spierfonds (neuromuscular fellowship grant W.F19-03).

